# Modelled Optimization of SARS-Cov-2 Vaccine Distribution: an Evaluation of Second Dose Deferral Spacing of 6, 12, and 24 weeks

**DOI:** 10.1101/2021.02.28.21252638

**Authors:** GT Jurgens, K Lackner

## Abstract

**Background:** Multiple recent studies have shown strong first dose vaccine efficacy for both Moderna mRNA-1273 and Pfizer/BioNTech BNT 162b2, which has stimulated discussion of maximizing initial population immunity during a time of vaccine shortage by using a deferred second dose strategy for these vaccines.

**Methods:** Our model examines the size of the effect of spacing of the second dose with 6, 12, and 24 week deferred spacing regimens relative to 3 week spacing.

**Results:** Deferring the second dose from 3 weeks to 6 weeks, 12 weeks, and 24 weeks shows progressive benefit to population immunity for any given time period, even with significant one dose efficacy decay. The benefits are influenced by vaccine supply per capita.

**Conclusion:** The longer the second dose is deferred the larger the benefit in initial population immunity, provided one dose efficacy does not significantly wane. Monitoring one dose efficacy duration from the UK or Quebec minimizes this risk, as the gathered data will help ensure the second dose is given at an optimal time. How this information is implemented should vary depending on the population and whether the goal is to optimally protect high risk groups or to increase total population immunity as quickly as possible. Benefits to deferring the second dose are influenced by the length of deferral, one dose efficacy, and vaccine supply per capita. The time to herd immunity could be shortened by 4 weeks with the implementation of a 12 week spacing regimen or 10 weeks with a 24 week spacing regimen.

## Background

The SARS-Cov-2 pandemic has inspired the creation of multiple effective vaccines, including a new type of vaccine using mRNA by Moderna (mRNA-1273) and Pfizer/BioNTech (BNT 162b2). Given the urgency of the pandemic and unknown immunogenicity of this new vaccine type, the manufacturers appeared to select doses at the highest tolerated level and spaced them with a short time frame to achieve a quick restimulation of the immune response. This sensible approach worked better than most would have anticipated, with a two dose efficacy rate of stopping symptomatic COVID at 94-95% in trial data (1,2), with reports from Israeli health agencies Maccabi and Cialit reporting similar figures in Israel’s population (3). Surprisingly, one dose efficacy against symptomatic disease was also very high 93-94% in trial data (2, 4) with multiple sources of data around the world confirming a 80% to 90% one dose efficacy once sufficient time is given for the immune system to mount a response (5, 6, 7). Lower viral titres in those who had received one dose that do get infected would point to decreased transmission of virus even without the second dose (8) and asymptomatic infections also appear to be reduced by 4 fold (9). Our previous model showed that even high estimates of decay rates for one dose efficacy were highly unlikely to nullify the benefit to population immunity that was gained by deferring the second dose (10). To the best of the authors’ knowledge, how this benefit changes with different spacing regimens as well as how it changes with vaccine availability has not yet been evaluated.

## Methods

We developed a model using a Python script to estimate the benefits to increased population immunity of deferring the second dose of the mRNA vaccines from 3 weeks to 6, 12, and 24 weeks. This modelling evaluates a population receiving vaccine shipments increasing by 10% per week such that the entire population can be fully immunized in 14 months. We also evaluated how to adjust these results for regions with higher or lower available vaccine supply per capita, given the size of the population and the available vaccine will vary by region. Decay rate of efficacy after 2 doses of the vaccine is set at 1% per month, and for one dose the decay rate is set at 2.5% per month, unless otherwise noted. A one dose efficacy of 93% and a two dose efficacy of 95% were used in our modelling, based on the trial data seen in Pfizer/BioNTech’s phase 3 data (1, 2). The total average population immunity was determined by calculating the area under the curve (unpublished data). Growth of the vaccine shipments were modelled at 10% increase per month. Immunity effects were realized two weeks after the respective doses were given.

A simple formula was developed to adjust for different observed efficacies as well as for populations that have partial natural immunity from COVID-19 infections.

## Results

Benefit to population immunity was seen in all spacing regimens longer than 3 weeks, whether examining a subset of the population (Fig. 1) or the entire population (Fig. 2). The benefits were proportionate to time the second dose was delayed; deferring the second dose to 12 weeks had approximately twice the benefit as deferring to 6 weeks, but only half the benefit of the 24 week regimen (Fig. 3, Fig. 4). Absolute population immunity gains of 2-20 percentage points were seen. These gains are increased significantly if the vaccine supply is more robust, see Figures 6-8. Viewed from another angle, a given population immunity level was reached from 1.5 to 10.5 weeks quicker by deferring the second dose. This benefit was diminished, but still present, even when using a high one dose decay estimation of 10% per month (Fig. 5).

**Figure 1:**
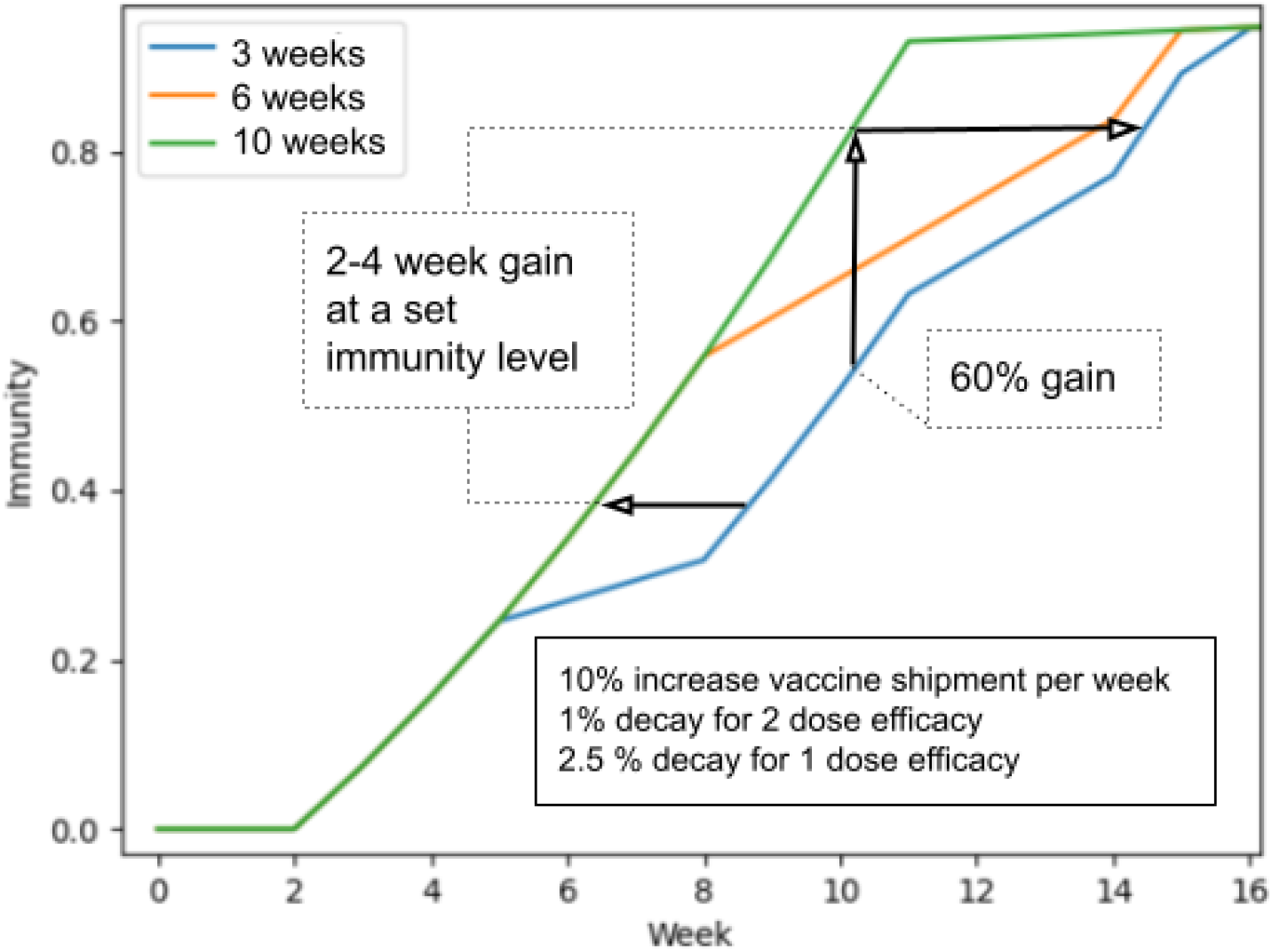
High Risk Population. Figure 1 shows an increased relative immunity of 60% at week 10 by deferring the second dose from 3 weeks to 10 weeks (absolute gain of 30 percentage points). It would take 4 more weeks for the standard 3 week spacing group to reach an equal immunity. The medium and low risk group profile were similar (unpublished data) 12 and 24 week spacing is not shown as all first doses were given by 10 weeks.

**Figure 2:**
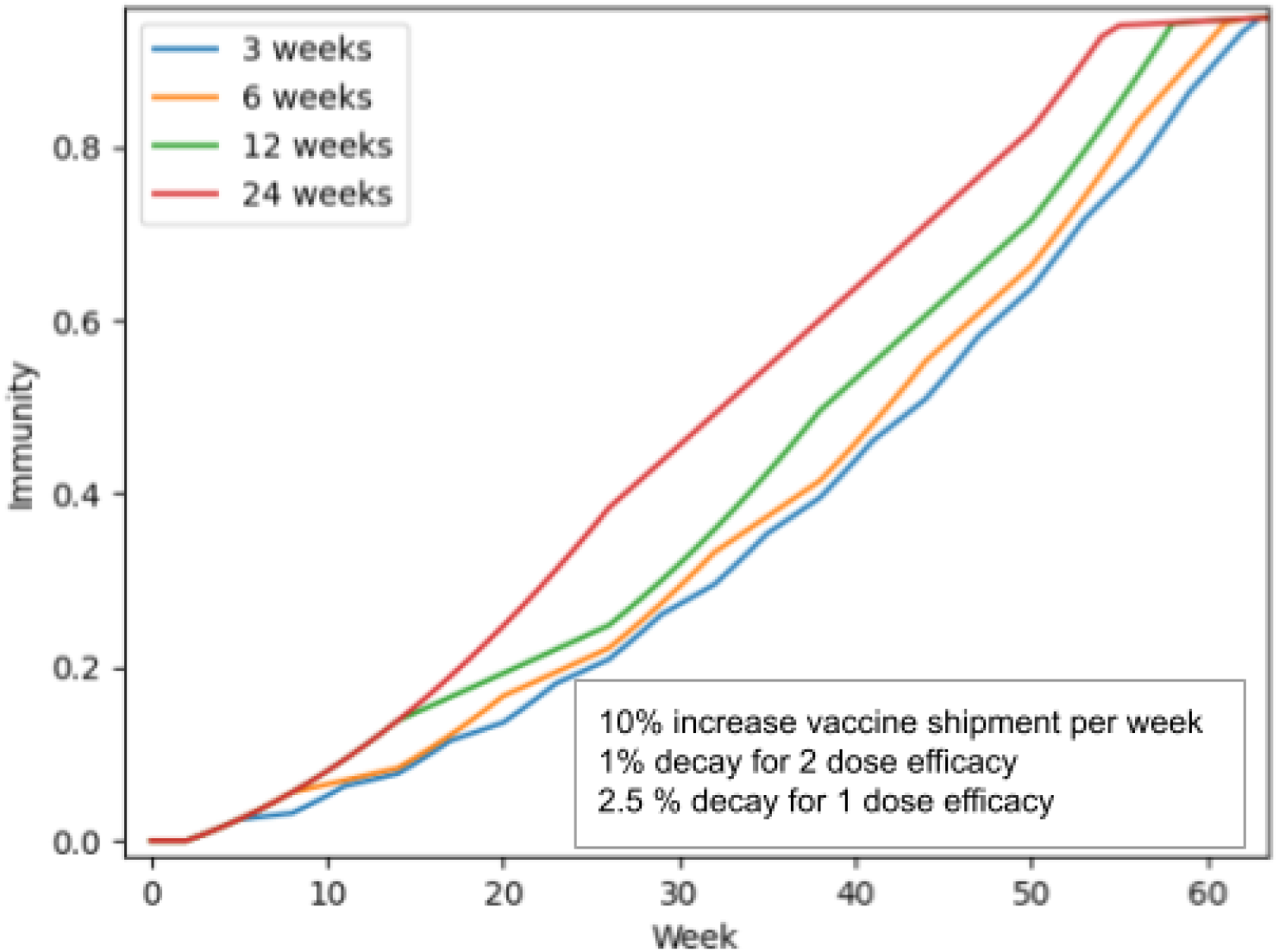
Entire Population Visualized. Figure 2 shows four different spacing regimens: the standard 3 week spacing is compared to 6, 12, and 24 week spacing. The longer the second dose can be deferred, me stronger the gain of population immunity at a given time, or faster the time period to reach a. given level of population immunity (as long as one dose efficacy does not dramatically wane).

**Figure 3A, 3B, 3C:**
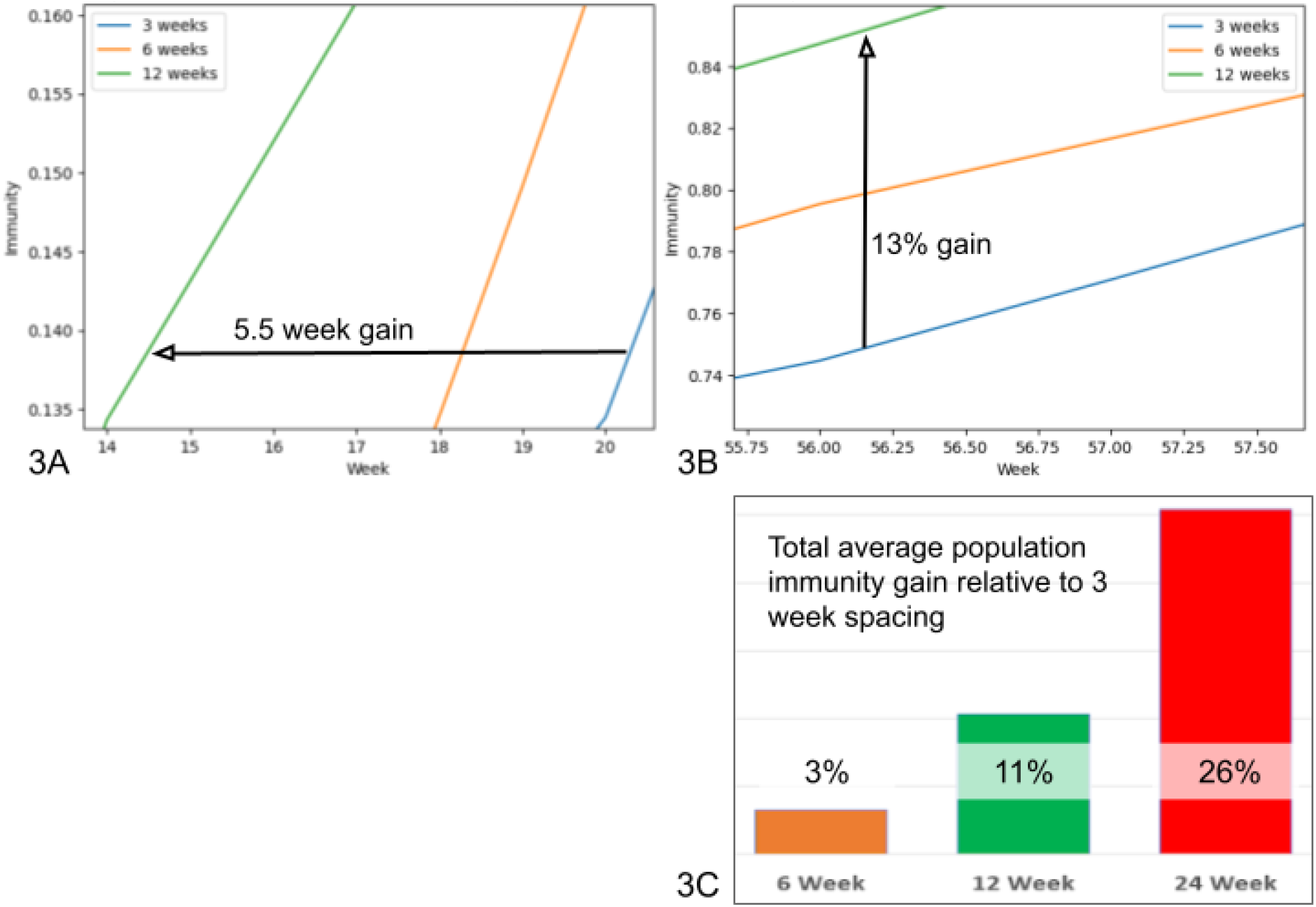
Evaluating 12 Week Spacing Benefits. Figure 3A: Benefits of deferring the second dose from 3 to 12 weeks can be shown by looking at how much faster a set level of immunity is obtained (in this case 5.5 weeks, see 3A) or the increase in population immunity at a set time (in this case a relative gain of 13%, or absolute gain of 10 percentage points, see 3B). Overall 12 week spacing reaches a set immunity level 4 to 6 weeks earlier than the 3 week spacing, and the relative gain in population immunity is 10%to 67%(an absolute gain of 4 to 10 percentage points).

**Figure 4:**
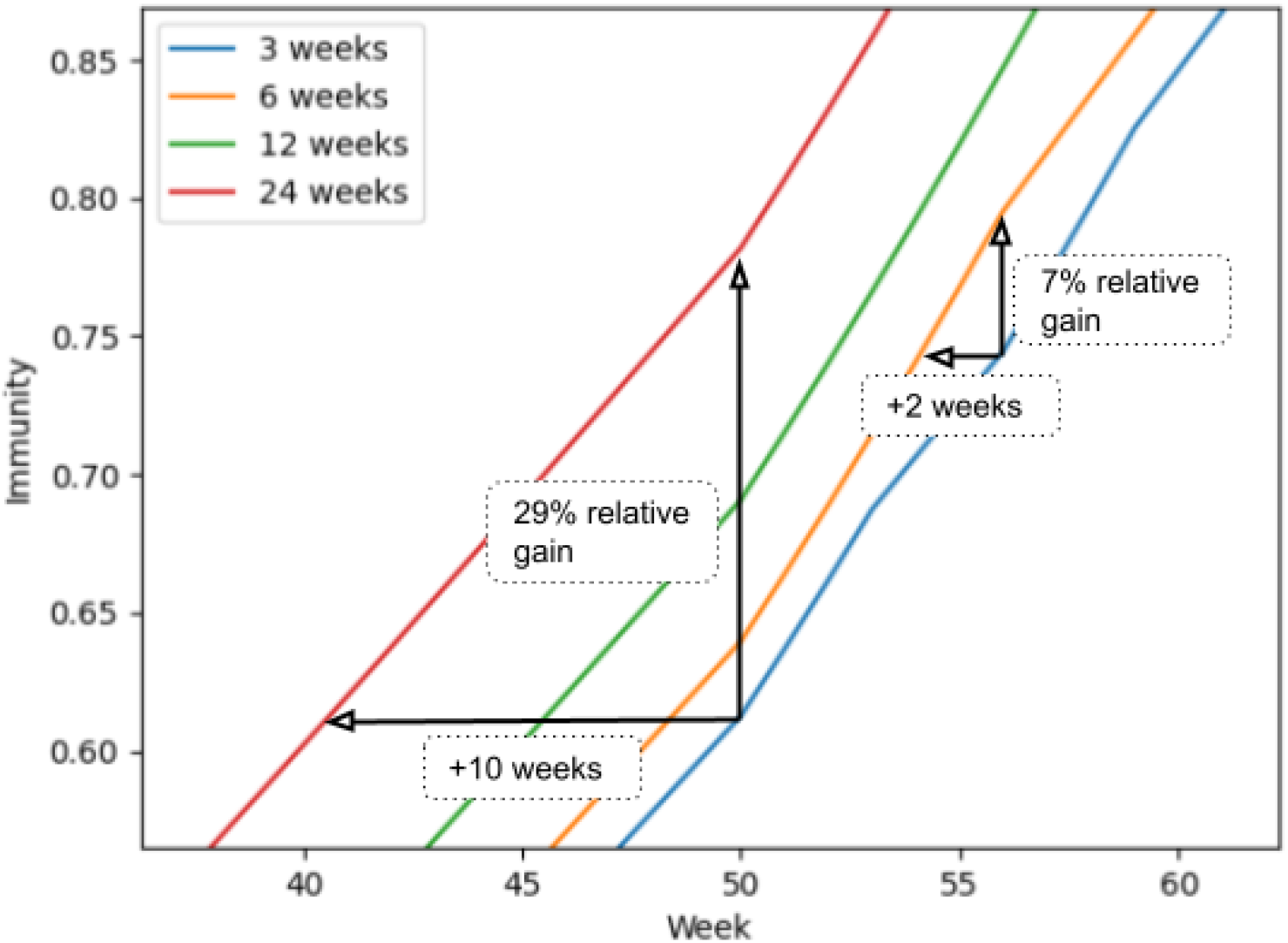
Evaluating 6 and 24 week spacing Benefits. Figure 4: 6 week spacing led to grains of 2 to 5 percentage points population immunity at set time, (relative increases change depending on the set time), and reached a set immunity level 1 to 2.5 weeks earlier. 12 week spacing led to gains of 10 to 20 percentage points population immunity at a set time, and reached a set immunity 8 to 10.5 weeks earlier.

**Figure 5:**
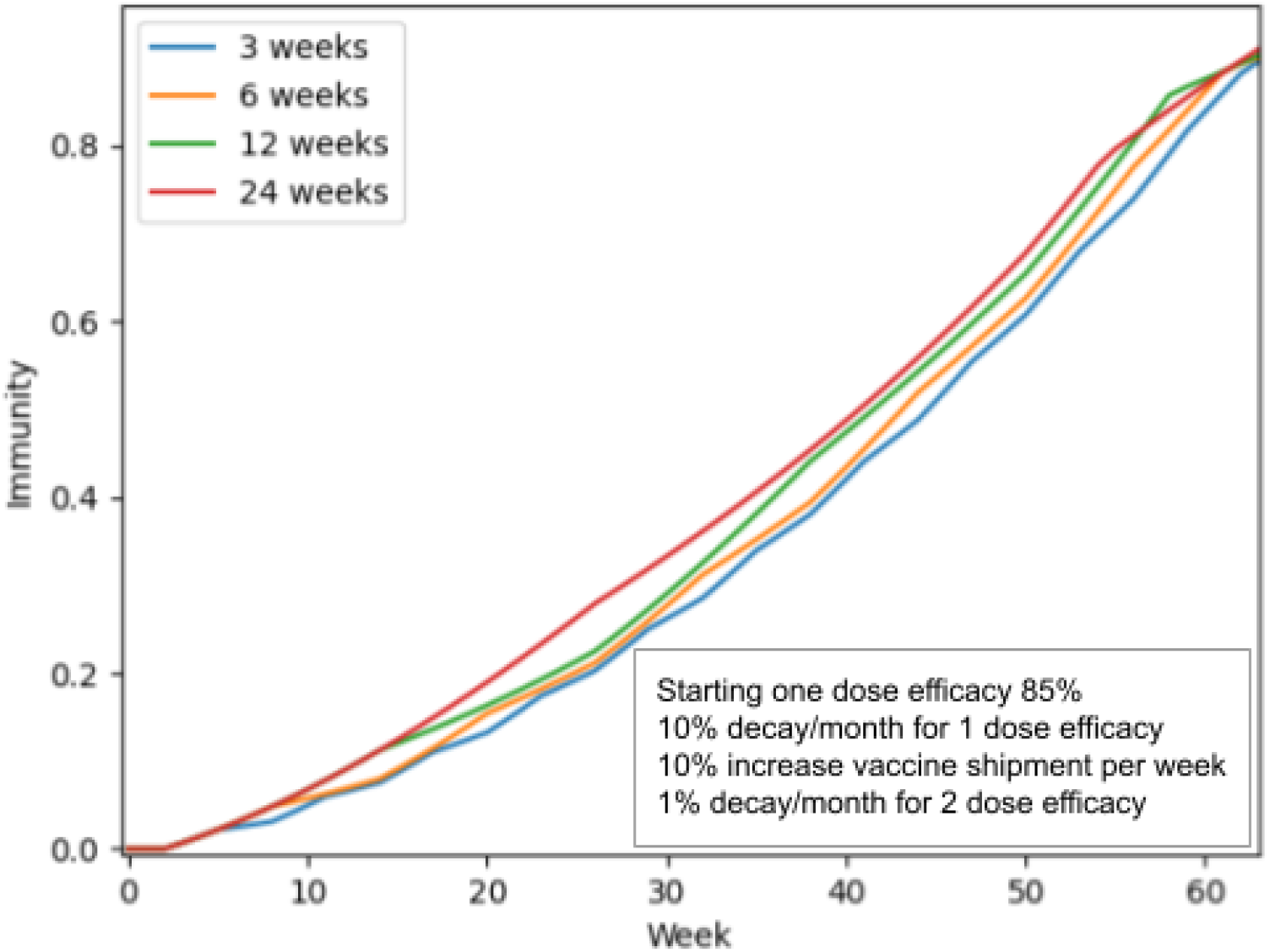
Rapid one Dose immunity Decay Evaluation. Figure 5: Even with very rapid one dose immunity decay of 10% pre month and estimated one dose efficacy of 85%, the overall population immunity is still benefited by deferring the second dose for all examined spacing durations

**Figure 6:**
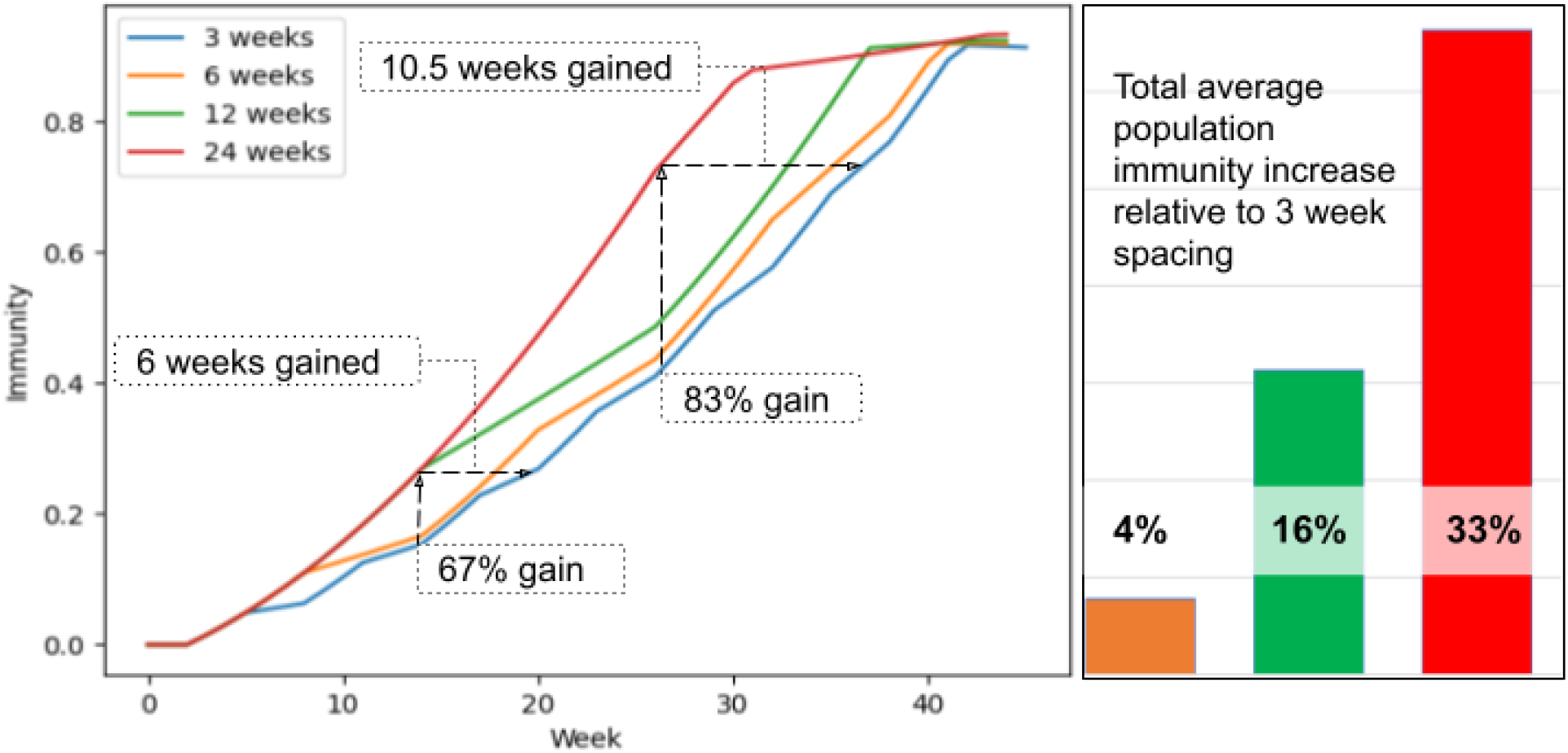
Increased Vaccine Supply Overview. Figure 6: A) For a vaccine supply that can fully vaccinate a population by 40 weeks, using a 24 week regimen increases population immunity by 83% relative to a 3 week regimen (an absolute increase of 33 percentage points) at 26 weeks. It would take another 10.5 weeks for a 3 week regimen to reach the equivalent level of immunity. For 12 week spacing, the relative immunity gain is as high as 67% at 14 weeks (absolute gain of 10 percentage points) and 6 weeks of time gained to reach that level of immunity. Right side: Average population immunity gain relative to 3 week spacing for the roll out period.

The estimated gains as modelled above are for a supply chain that can supply enough doses to fully vaccinate the population in about 60 weeks. That is a conservative estimate of some regions’ supply chains, and examining how supply will change these benefits can be useful in adapting this modelling to a region of interest. Changing the vaccine supply per capita does change the benefit of increased population immunity at a given time differently than the benefit of reaching a set population immunity sooner, see Table 1 and Fig. 6. If a region has a more robust vaccine supply per capita so that they can fully vaccinate their population within 40 weeks, that will modify the magnitude of benefit modelled. Namely, this increases the gain in population immunity, but the gain in time to reach a set immunity is relatively unchanged. Alternatively, for regions expecting to take longer than a year to vaccinate their population, lower gains with population immunity would be expected if the same spacing regimen was used.

**Table 1:** Magnitude of Benefits Depend on Vaccine Supply Per Capita

| Vaccine supply per capita | Benefit of increased population immunity at time $x$ | Benefit of immunity level $y$ being reached sooner |
| --- | --- | --- |
| High | +++++ | +++ |
| Average | +++ | +++ |
| Low | ++ | +++ |

The influence of vaccine supply can be illustrated by comparing the 24 week spacing gains shown in Figure 7, which shows a population with robust vaccine supply per capita, as well as medium supply, and low supply. The more robust the vaccine supply, the greater the increase of population immunity for a 12 week spacing compared to a 3 week spacing regimen. The 24 week spacing compared to the 3 week spacing regimen illustrates a gain of 78% for robust vaccine supply, 58% for medium vaccine supply, and 26% for low supply. These correspond to absolute population immunity percentage point increases of 32, 19, and 15, respectively. With a very robust vaccine supply, a 24 week spacing regimen is not needed as the entire population will be able to receive their first dose 24 weeks has passed, see Figure 8.

**Figure 7:**
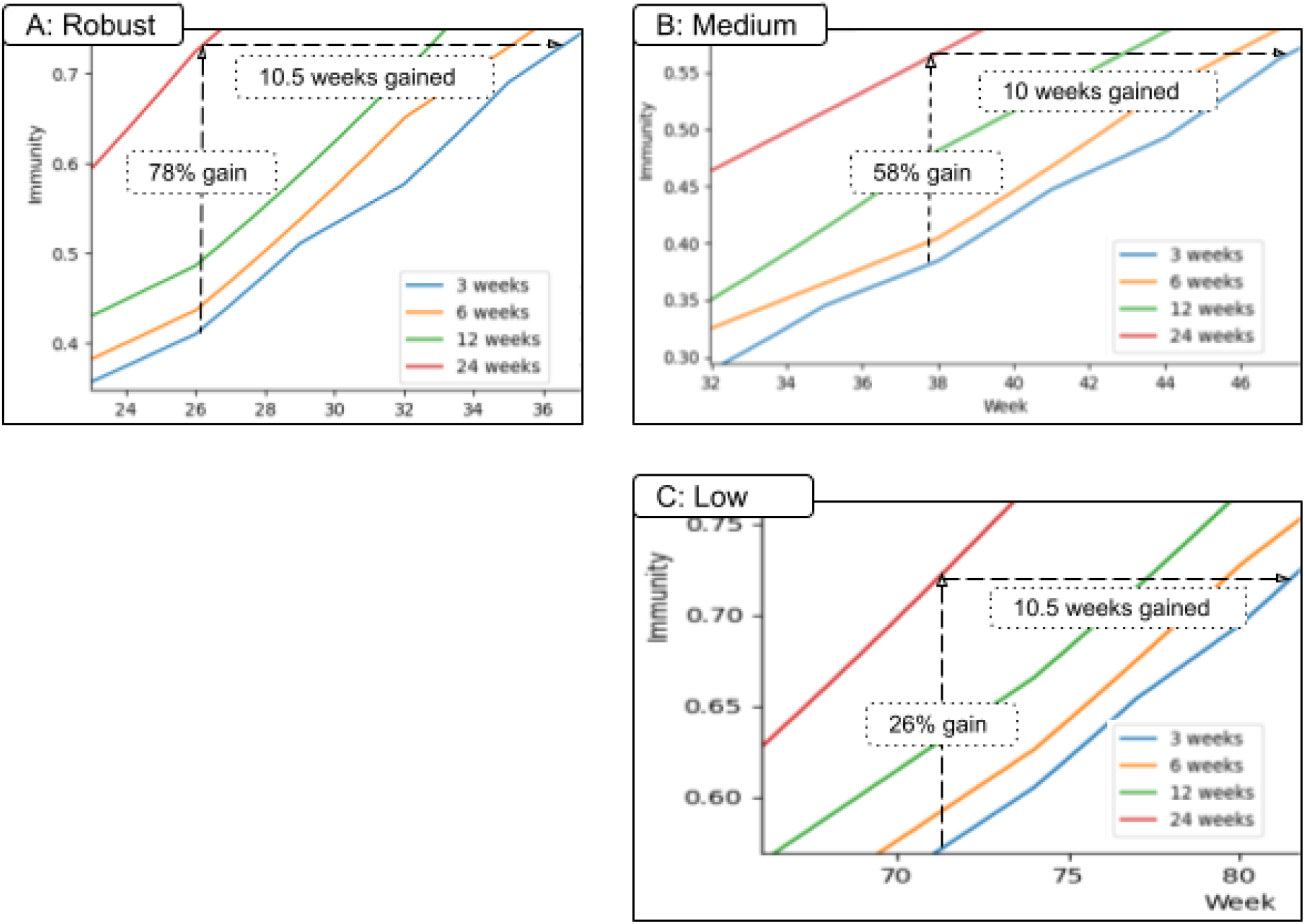
Different Vaccine Supply Production Influences Spacing Benefits. Figure 7 compares robust, medium, and low vaccine supply chain, A-C respectively. A: By the 26th week, the 24-week-spacing protocol results in population immunity of 78% higher than the 3-week-spacing (an absolute increase of 32 percentage points). B: 58% relative immunity gain is observed, an absolute increase of 19 percentage points. C: 26% relative gain is seen, an absolute increase of 15 percentage points. For each of these gains, note that it would take at least another 10 weeks for the 3 - week-spacing protocol to catch up and reach the same level of immunity.

**Figure 8:**
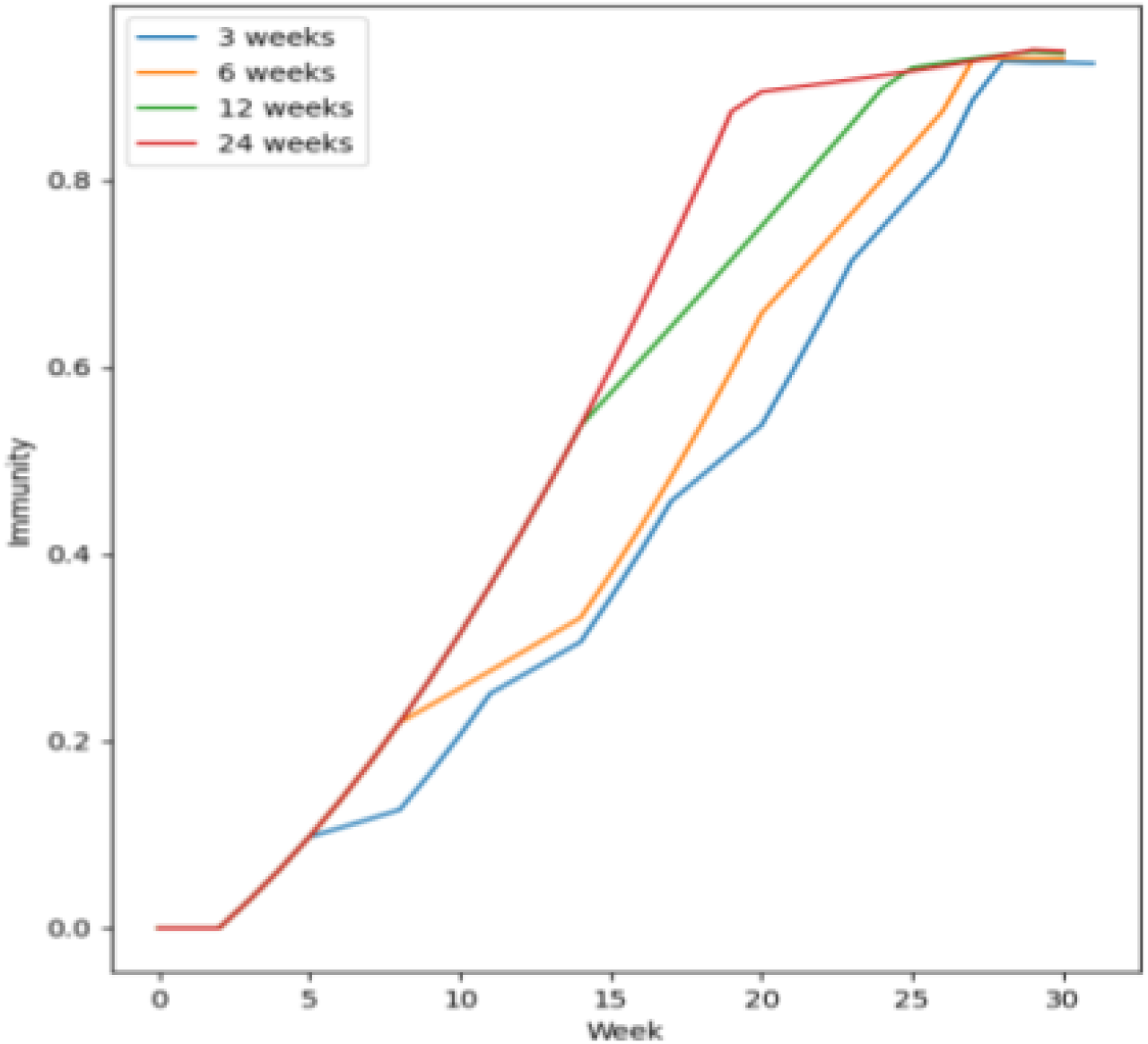
Very Robust Vaccine Supply. Figure 8: The optimal vaccine distribution is to defer all second doses until everyone has had their first dose. This is visualized here, with a very robust vaccine supply ensuring everyone has received their first vaccine dose by ∼16 weeks. There is no advantage to spacing the doses out further. The gains over 6 and 12 week spacing area readily apparent

Gain in immunity is a function of the spacing used relative to the total roll out time. 24 week spacing makes up a much larger share of a 40 week total timeline than it does of an 80 week total timeline and thus results in higher relative immunity gains. If a longer roll out is required due to less robust vaccine supply, a longer spacing protocol proportionate to the roll-out time period would increase the relative gains. Of course, such longer spacing would come with the appropriate caveats of waning one dose efficacy considered in this paper.

Conversely, notice that the gain in time to reach a set immunity level appears only minimally affected by vaccine supply, if at all. How much faster a set immunity can be reached compared to 3 week spacing is primarily determined on the spacing regimen used: 24 week spacing gains ∼10.5 weeks, 12 week spacing gains ∼4.5 weeks, and 6 week spacing gains ∼1.5 weeks. The longer that the second dose is deferred, the greater the time saved before reaching a set immunity point.

As one would expect, the benefit observed in the model decreased proportionately with decreased one dose efficacy, such that no benefit is seen in deferring the second dose when the first dose efficacy is half of two dose efficacy. Our model uses the trial data of 93% efficacy after one dose and 95% after two doses, thus the net benefit to diverting a second dose to use as a first dose instead is 91 percentage points (93-(95-93)= 91). The modelled gains can also be adjusted for differing one dose and two dose efficacy rates for the population of interest using the following formula:

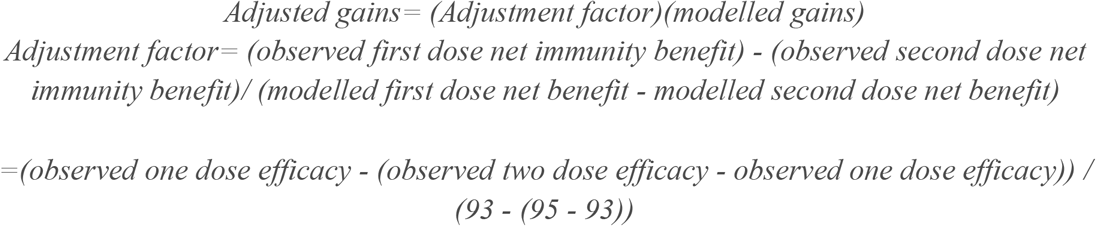

For example, with 72% one dose efficacy and 86% two dose efficacy against symptomatic and asymptomatic infection as reported in the SIREN study (11), the above formula gives an adjustment factor of 0.64, meaning the benefits to deferring the second dose would be about a third less than currently modelled.

The above formula can also be used to adjust for vaccinating the cohort who have recovered from COVID-19. Previous infection has been estimated to reduce risk of a subsequent infection by 94% for a period of at least 5 months (12). Vaccinating these recovered individuals only increases population immunity by likely 2 or 3 percentage points (to a maximum of 6 percentage points), which is a small gain in immunity compared to the 93 percentage points gained by vaccinating a naive individual. For each *x*% of the doses going to the COVID-19 recovered cohort (rather than naive individuals), the expected benefit of the first dose falls by nearly *x*% as well. If 10% of the population being vaccinated already has immunity, the expected gains as modelled would decrease by nearly 10%.

## Conclusions

The modelling above demonstrates a clear benefit to population immunity with all second dose deferred regimens, and second dose deferral maintains this advantage even when one dose decay is an order of magnitude larger than two dose efficacy decay, as our previous paper also showed (10). By evaluating the time to reach a set immunity level within the entire population (rather than solely focusing on the population immunity gains at a set time), the above modelling shows that time to reach herd immunity could be shortened by 1.5 weeks with a 6 week spacing regime, 4.5 weeks with 12 week spacing, and 10.5 weeks with 24 week spacing, independent of vaccine supply.

A few additional insights have been illustrated by modelling different spacing regimens and varying vaccine supply scenarios. First, the longer the spacing regimen is used, the more benefit is seen in population immunity and in shortening the time needed to reach a given set immunity point. This benefit is proportionate, with a 12 week spacing conferring twice the benefit of a 6 week spacing regimen relative to the standard 3 weeks. Furthermore, the gains for the deferred second dose accumulate throughout the roll out period - resulting in a higher overall average population immunity week by week for every deferral.

Second, vaccine supply does not impact the gains made in shortening the time to reach a certain set point, but it does impact population immunity levels, with more robust vaccine supply leading to increased population immunity.

Third, vaccinating previously recovered individuals who have a high natural immunity to COVID-19 is inefficient. There are multiple reasons why the COVID-19 recovered cohort might get vaccinated, including maximizing individual immunity for those at high risk and inadvertently vaccinating those who had asymptomatic infections. However, deferring the first dose of vaccine for this cohort until everyone else has received their first dose is optimal from a population immunity perspective, a factor that public health organizations should consider when implementing a distribution plan.

Fourth, as increased benefits were noted with increased spacing, the optimal way to increase population immunity is to defer the second dose until everyone has received their first, as long as one dose efficacy does not rapidly wane.

A common concern about implementing the deferred second dose strategy is that the one dose efficacy duration is unknown for now. AstraZeneca’s one dose trial data showed sustained one dose immunity for at least 12 weeks (13), but given that it did not use the novel mRNA vaccine technology, the concern around one dose duration was still valid. However, with multiple ongoing studies evaluating one dose efficacy duration of the mRNA vaccines (such as in Quebec and the UK), this risk has greatly been diminished. Any region that now implements a second dose deferred strategy - weeks after these other trials have started - will have sufficient notice of when one dose efficacy decays, as long as they monitor these other public health authorities’ data. In other words, the duration of one dose efficacy for the mRNA vaccines is still not known, but it will be known soon, and should give ample leeway to inform the optimal time to give the second dose for this new mRNA vaccine technology.

How dose deferral is implemented will depend on the characteristics of the population in question. One strategy would be to defer the second dose only until all high risk individuals have first received their first dose, and then the second dose is given as vaccine supply allows. Once the high risk group has received both doses, the same distribution strategy is implemented for the medium and low risk groups, respectively. Given that the elderly may have a level of immune senescence leading to a delayed response to the vaccine (14), with possibly a lower one dose efficacy than the initial trial data suggested (15), this may be a prudent option. This appears to be the strategy Quebec is taking, as it optimizes immunity for the most vulnerable (6), and focusing vaccination on the elderly appears to prevent the most deaths as well as minimizing total expected years lost (16).

An alternate strategy may be to defer the second dose as long as one dose efficacy does not appear to significantly wane, in such a way that the entire population would receive their first dose before anyone gets a booster. This strategy would lead to an optimal increase in population immunity but may leave the highest risk groups vulnerable with lower one dose efficacy levels.

Another valid concern with deferred second dosing is that very prolonged immune spacing might also diminish the booster effect of a second dose. Individuals who have been infected with COVID-19 have shown dramatic and robust responses to a single dose of a mRNA vaccine even 10 months after recovery, even in people whose antibodies have waned to unmeasurable levels (17). Furthermore, AstraZeneca’s data showed improved immunity with a 12 week spacing window compared to less than 6 week spacing (13). This helps allay this concern to a certain degree, but the optimal spacing for a booster dose may differ after infection or based on using different vaccine technology. A reasonable way to evaluate this might be to give a second dose to a small sample of the population every week during the deferral period and ensure they have large increases in their immune markers consistent with a boost response. When (or if) the immune markers stop having a robust increase as expected, then that would help determine the maximal spacing to optimally restimulate the immune system.

With studies from multiple sources now confirming strong one dose efficacy, and with studies in progress of one dose efficacy duration underway, the risk to implementing a second dose deferral strategy has been minimized. Multiple previous models have shown benefit to deferring the second dose (10, 18, 19). The modelling done here helps characterize the benefits to deferring the second dose, adding to the growing evidence that continuing with the standard 3 week spacing regimen will likely result in preventable mortality and morbidity.

## Data Availability

The datasets generated during and/or analysed during the current study are available from the corresponding author on reasonable request.

